# A Machine Learning-Based Prediction of Hospital Mortality in Mechanically Ventilated ICU Patients

**DOI:** 10.1101/2024.07.12.24310325

**Authors:** Hexin Li, Negin Ashrafi, Chris Kang, Guanlan Zhao, Yubing Chen, Maryam Pishgar

## Abstract

**Background:** Mechanical ventilation (MV) is vital for critically ill ICU patients but carries significant mortality risks. This study aims to develop a predictive model to estimate hospital mortality among MV patients, utilizing comprehensive health data to assist ICU physicians with early-stage alerts.

**Methods:** We developed a Machine Learning (ML) framework to predict hospital mortality in ICU patients receiving MV. Using the MIMIC-III database, we identified 25,202 eligible patients through ICD-9 codes. We employed backward elimination and the Lasso method, selecting 32 features based on clinical insights and literature. Data preprocessing included eliminating columns with over 90% missing data and using mean imputation for the remaining missing values. To address class imbalance, we used the Synthetic Minority Over-sampling Technique (SMOTE). We evaluated several ML models, including CatBoost, XGBoost, Decision Tree, Random Forest, Support Vector Machine (SVM), K-Nearest Neighbors (KNN), and Logistic Regression, using a 70/30 train-test split. The CatBoost model was chosen for its superior performance in terms of accuracy, precision, recall, F1-score, AUROC metrics, and calibration plots.

**Results:** The study involved a cohort of 25,202 patients on MV. The CatBoost model attained an AUROC of 0.862, an increase from an initial AUROC of 0.821, which was the best reported in the literature. It also demonstrated an accuracy of 0.789, an F1-score of 0.747, and better calibration, outperforming other models. These improvements are due to systematic feature selection and the robust gradient boosting architecture of CatBoost.

**Conclusion:** The preprocessing methodology significantly reduced the number of relevant features, simplifying computational processes, and identified critical features previously overlooked. Integrating these features and tuning the parameters, our model demonstrated strong generalization to unseen data. This highlights the potential of ML as a crucial tool in ICUs, enhancing resource allocation and providing more personalized interventions for MV patients.

## Introduction

In the United States, over one million patients receive mechanical ventilation (MV) annually in Intensive Care Units (ICU), occupying 24–41% of ICU beds at any given time [1]. Although MV is frequently considered a lifesaving intervention, patients undergoing non-surgical procedures that require MV have a hospital mortality rate exceeding 35% [2].

This study aims to develop an improved predictive model to estimate the mortality of MV patients using patient health data, including early-stage symptoms [3]. The model is intended to assist ICU physicians in early alerting by utilizing a comprehensive database containing easily obtainable and well-generalized health data for better accuracy and prediction [4]. Previous research has demonstrated the feasibility of using patient health data for predictive modeling in ICU settings, highlighting the importance of integrating such models into clinical practice [5, 6].

There are many factors to consider when assessing risk for mechanically ventilated patients, including predictors available on the first day, which can be used to predict hospital mortality [7]. Effective machine learning prediction requires careful feature selection, accounting for the severity of the disease and outcomes to understand early prediction factors and the impact of MV [8, 9]. Previous studies have shown that XGBoost performs well in predicting hospital mortality, aiding in understanding different clinical situations for early-stage alerting [10, 11]. However, different models may be suitable for various purposes beyond performance, considering clinical outcomes and contributing factors, which can influence mortality rates [12, 13].

From the present study, CatBoost performed better than other approaches. Clinical situations and feature selection are crucial in assessing both short- and long-term factors [14–16]. By implementing machine learning approaches, we can better understand how these methods, combined with current healthcare data methodologies, improve the evaluation of risk factors [17, 18].

We thoroughly examined the inclusion criteria for our feature selection process using the MIMIC-III database when assessing risk factors, followed by various data extraction techniques for data preprocessing. Additionally, we implemented several machine learning models, each yielding unique results and offering different perspectives on predicting the mortality rate for MV patients.

## Methadology

### Data availability

The Medical Information Mart for Intensive Care III (MIMIC-III) is a publicly accessible dataset [4]. It contains de-identified health information from over 40,000 ICU admissions at the Beth Israel Deaconess Medical Center, covering the years 2001 to 2012 [19]. Developed by the MIT Lab for Computational Physiology, MIMIC-III includes a wide range of data categories, such as demographics, vital signs, laboratory test results, medications, and mortality outcomes. This comprehensive dataset supports extensive research in clinical informatics.

### Patient selection

We initially included patients who required mechanical ventilation (MV) in the ICU. To exclude incomplete and duplicated data, we refined the dataset according to specific inclusion criteria: (I) patients aged between 18 and 90 years; (II) patients with complete mortality information; (III) patients with sufficient clinical data, ensuring columns with fewer than 90% missing data were included. We utilized ICD-9 codes to identify relevant patient records and linked these with ventilation-related data, resulting in a final cohort of 25,202 patient samples. The flowchart of patient selection and data preprocessing is illustrated in Fig 1.

**Fig 1.**
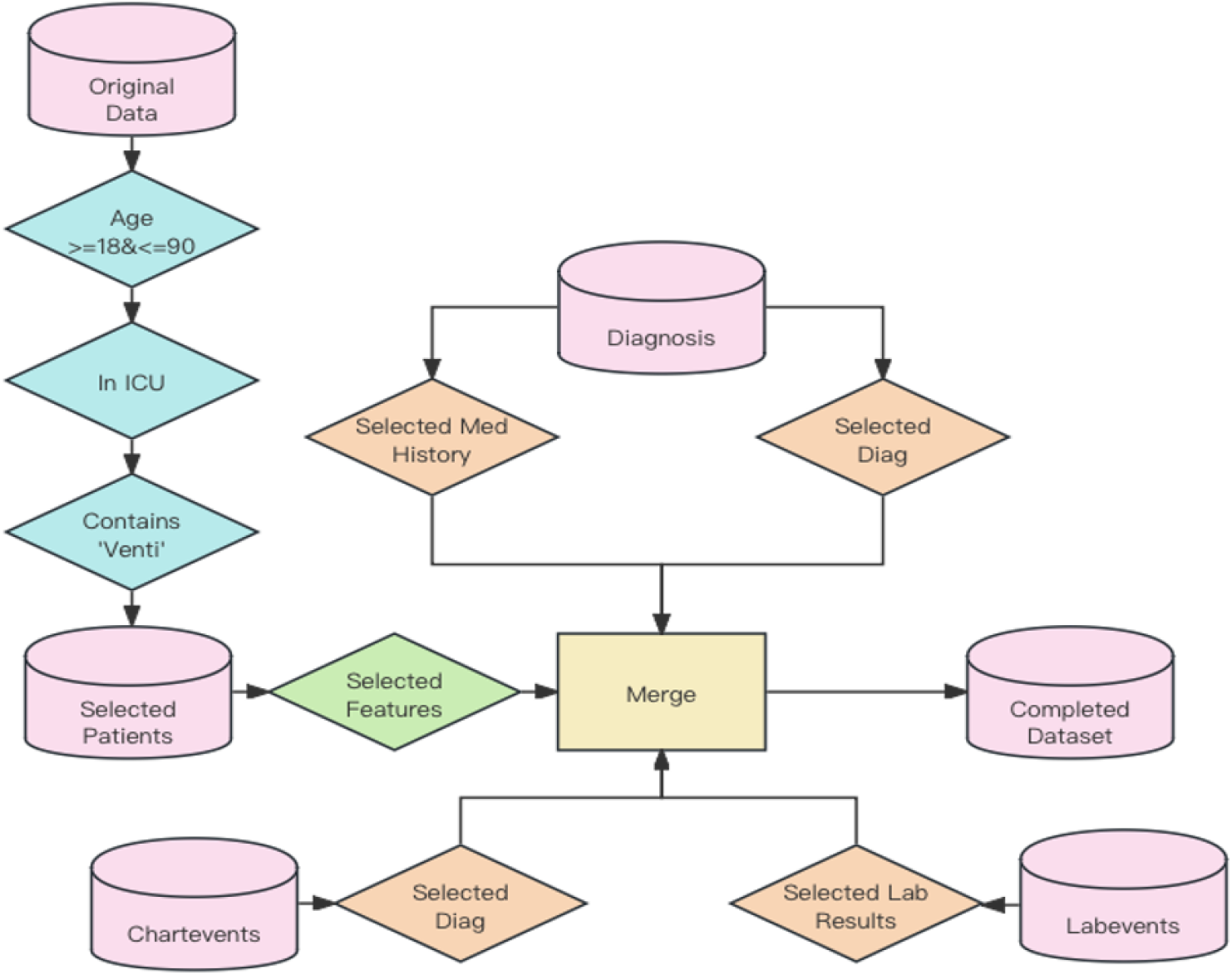
Patient Selection. Flowchart illustrating the inclusion criteria and data preprocessing steps leading to the final cohort of 25,202 patient samples.

### Feature selection and data preprocessing

The feature selection process in this study involved several stages. Initially, backward elimination and the Lasso method were employed to identify the most significant features [20, 21]. This selection was further refined through an extensive review of existing literature and clinical insights, resulting in the selection of 32 features.

The demographic data included age. Vital signs such as heart rate (HR), respiratory rate (RR), respiratory rate set (RR Set), temperature (TEMP), non-invasive blood pressure systolic (NIBP Systolic), non-invasive blood pressure diastolic (NIBP Diastolic), arterial blood pressure systolic (ABP Systolic), and arterial blood pressure diastolic (ABP Diastolic) were recorded. Additionally, laboratory values encompassing bicarbonate, serum creatinine, serum potassium concentration, and serum sodium concentration were included. Comorbidities such as liver failure, chronic heart failure, organ failure, sepsis, uncomplicated hypertension, and respiratory dysfunction were also documented. Differences in vital signs, such as the differences in heart rate, respiratory rate, NIBP systolic, and temperature, were calculated. The detailed overview of feature information used in this study is presented in Table 1.

**Table 1.** Detailed Overview of Feature Information.

| Feature Type | Feature Name | Feature Type | Feature Name |
| --- | --- | --- | --- |
| Lab Results | Bicarbonate_max_value | Vitals | Heart Rate_min |
|  | Bicarbonate_min_value |  | Respiratory Rate_min |
|  | Serum_Creatinine_min_value |  | Respiratory Rate Set_min |
|  | Serum_Creatinine_diff |  | Temperature C (calc)_min |
|  | Serum_Potassium_Concentration_max_value |  | Arterial BP [Diastolic]_min |
|  | Serum_Sodium_Concentration_min_value |  | Heart Rate_max |
|  | Serum_Sodium_Concentration_diff |  | Non Invasive Blood Pressure systolic_min |
| Demographics | Age |  | Non Invasive Blood Pressure diastolic_min |
| Comorbidities | liver_failure_count |  | Temperature Fahrenheit_min |
|  | Chronic_heart_failure_count |  | Arterial BP [Systolic]_max |
|  | organ_fail_count |  | NBP [Systolic]_max |
|  | sepsis_count |  | Respiratory Rate Set_max |
|  | Uncomplicated_hypertension_count |  | Arterial BP [Diastolic]_max |
|  | respiratory_dysfunction_count |  | Temperature F_max |
| Differences | Heart Rate_diff |  |  |
|  | Respiratory Rate_diff |  |  |
|  | NBP [Systolic]_diff |  |  |
|  | Temperature F_diff |  |  |

To address class imbalances in the target variables, the Synthetic Minority Over-sampling Technique (SMOTE) was applied [22]. Data preprocessing involved removing columns with over 90% missing data and applying mean imputation to the remaining missing values. Categorical features were converted into numerical codes using a Label Encoder to integrate them into regression and machine learning models. These preprocessing steps were essential for standardizing the dataset, enabling efficient model training and evaluation, and ensuring that analyses accurately reflected the original measurements and categories present in the dataset.

### Model development and optimization

Our final dataset included 25,202 patients with 32 features. We performed a 70/30 train-test split to facilitate model evaluation, choosing this method over cross-validation to improve performance and computational efficiency. We developed and assessed several machine learning (ML) algorithms, including Logistic Regression, CatBoost, XGBoost, Decision Tree, Random Forest, Support Vector Machine (SVM), and K-Nearest Neighbors (KNN).

The performance of these models was primarily measured using the Area Under the Receiver Operating Characteristic (AUROC) scores, with accuracy and F1 scores also calculated for a comprehensive comparison. Given the widespread use of AUROC in existing literature, it was selected as the primary metric for model evaluation [23]. CatBoost emerged as the top performer, confirming its superior performance relative to other models. Introduced in 2017, CatBoost is a novel boosting method based on Gradient-Boosted Decision Trees (GBDT). It offers several advantages, including support for categorical variables, streamlined parameterization, fast prediction capabilities, and notable accuracy [24]. Our study highlights the advantages of using CatBoost in hospital mortality prediction.

To provide a thorough comparison, we included six widely employed machine learning algorithms as baseline models: Decision Tree, Random Forest, SVM, KNN, Logistic Regression, and XGBoost. Decision trees partition the feature space hierarchically, minimizing impurity in each split [25]. Random Forest, an ensemble method, aggregates multiple decision trees for prediction [26]. SVM seeks an optimal hyperplane for class separation, maximizing the margin [27]. KNN assigns class labels based on the majority vote among the k nearest neighbors [28]. Logistic Regression estimates binary outcome probabilities using a logistic function [29]. XGBoost, an accelerated gradient boosting implementation, iteratively enhances predictive accuracy. These diverse methodologies allowed us to comprehensively evaluate CatBoost’s performance [30].

In comparing CatBoost with these baseline models, distinct differences emerged [31]. CatBoost’s symmetric algorithm for gradient-boosted decision trees provided significant enhancements in accuracy, robustness, and computational efficiency [32]. Evaluation metrics such as accuracy, precision, recall, F1-score, AUROC metrics, and calibration plots highlighted the predictive strengths of each model across various criteria. By employing these methodologies, we developed a robust predictive model for hospital mortality in mechanically ventilated ICU patients. This model offers valuable insights, supporting clinical decision-making and optimizing resource allocation within ICUs.

### Statistical analysis of models

To validate the statistical robustness of our model results, we conducted a comprehensive statistical analysis comparing the train and test sets using t-tests and chi-square tests. The dataset was split into a 70% train set and a 30% test set, which were used to evaluate the performance of trained models. A threshold p-value of 0.05 was set to determine the significance of differences [33]. This analysis focused on identifying any significant discrepancies between the two datasets, ensuring the reliability of the study’s findings.

T-tests were employed to compare the means of continuous variables between the train and test sets. For categorical variables, chi-square tests were utilized to assess the independence between the two sets. We tested the hypothesis that there was no significant difference between the train and test sets. If the p-value for each variable was greater than 0.05, we could reject the null hypothesis and conclude that the test set had notable distinctions from the train set. This analysis was crucial for confirming the consistency and generalizability of our model.

### Features importance

We analyzed each variable’s impact on our model using SHAP (SHapley Additive exPlanations) values, which measure feature importance. SHAP values reveal the most influential features in predicting the model’s output by ranking them based on their impact [34].

The graphical representation of SHAP values includes a bar plot of mean absolute SHAP values, indicating how much each feature contributes to the model’s predictions.

For instance, ‘Age’ was identified as the most significant feature, followed by ‘Bicarbonate max value’ and ‘respiratory dysfunction count’. These top-ranked features had higher mean absolute SHAP values, showing their significant impact on the model’s output. Lesser-impact features, such as ‘Respiratory Rate min’ and ‘Temperature F max’, had lower SHAP values. The SHAP summary plot, presented in Fig 2, visually demonstrates these findings. This analysis provides valuable insights into the internal mechanics of our machine learning models, ensuring transparency, improving model accuracy, and informing decision-making by highlighting key drivers of predictions.

**Fig 2.**
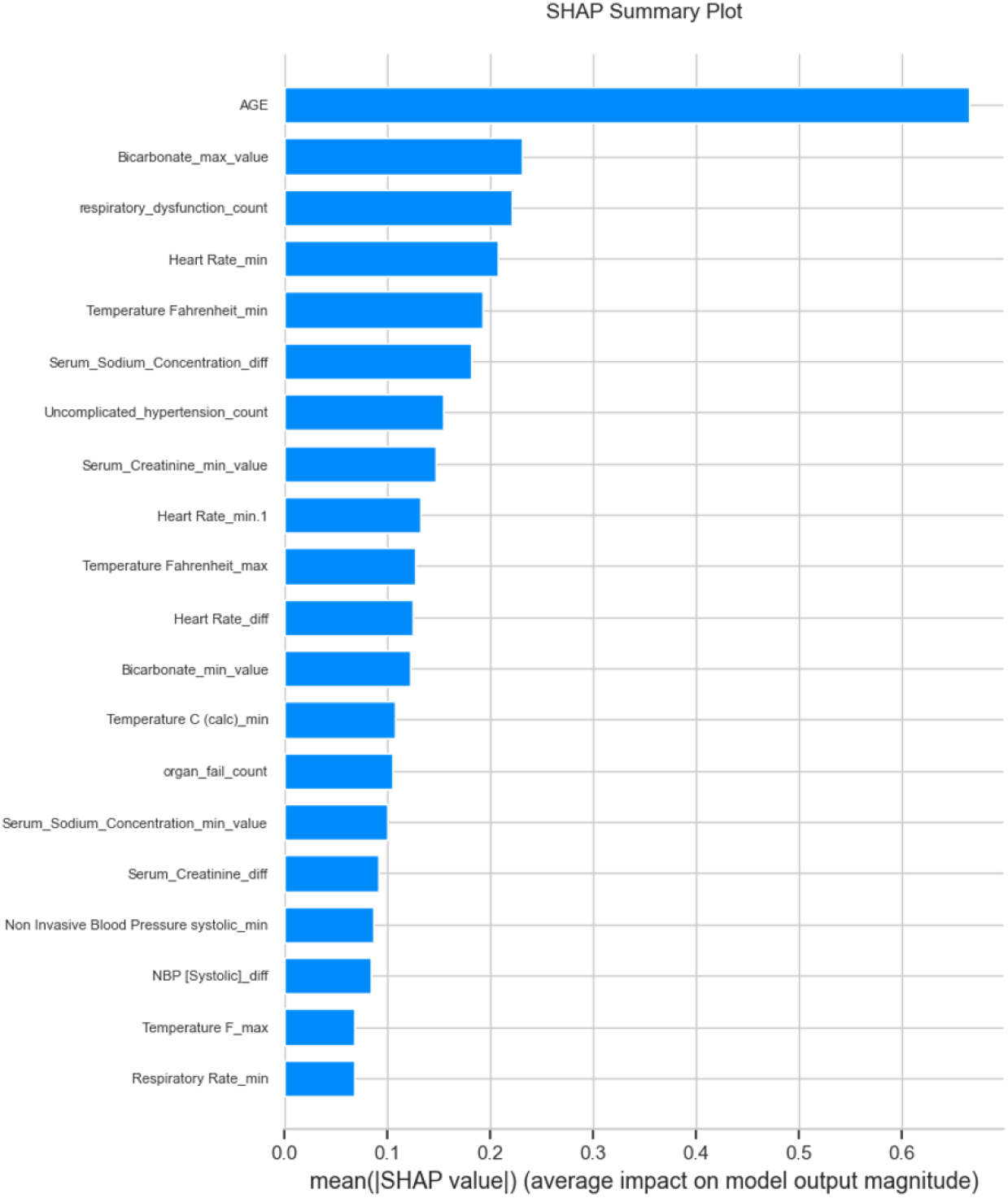
SHAP Analysis. Bar plot of mean absolute SHAP values, demonstrating the impact of each feature on the model’s predictions, with ‘Age’ being the most significant, followed by ‘Bicarbonate max value’ and ‘respiratory dysfunction count’.

This graphical representation provides a valuable tool for understanding the inner workings of complex machine learning models. By interpreting the average impact of each feature on model output magnitude, we gain insights into which features drive predictions the most and help ensure transparency in the predictive process. This, in turn, aids in debugging, improving model accuracy, and making informed decisions. This analysis provides valuable insights into the internal mechanics of our machine learning models, ensuring transparency, improving model accuracy, and informing decision-making by highlighting key drivers of predictions.

## Results

### Cohort characteristics model completion

Following our approach for feature selection and data preprocessing for ICU patients requiring mechanical ventilation, our final dataset included 25,202 patients from the MIMIC-III database. The selected cohort was randomly split into training and testing sets with a ratio of 70/30, resulting in 17,641 patients for training and 7,561 patients for testing. The training set was used to train the models, while the testing set was employed to evaluate the performance of our proposed model. To rigorously evaluate the disparity between the training and testing sets, we conducted a statistical analysis focusing on the hypothesis that the testing set exhibited notable distinctions from the training set. A threshold p-value of 0.05 was set to ascertain the significance of the differences. The analysis aimed to identify any substantial gaps between the two datasets, ensuring the reliability of the study’s findings. The detailed cohort values and p-values reflecting differences between the training and testing sets are presented in Table 2.

**Table 2.** Detailed Overview of Cohort Characteristics for train and test cohort. Values are presented as means with the standard deviations in parentheses.

| Characteristics | Train Cohort (N=17,638) | Test Cohort (N=7,562) | T-Stat | P-Value |
| --- | --- | --- | --- | --- |
| Age | 63.12(16.50) | 62.97(16.41) | 0.68 | 0.49 |
| Heart Rate_min | 60.02(16.77) | 60.07(16.79) | -0.2 | 0.84 |
| Respiratory Rate_min | 8.31(4.23) | 8.35(4.21) | -0.6 | 0.55 |
| Respiratory Rate Set_min | 10.12(3.72) | 10.07(3.76) | 1.18 | 0.24 |
| Temperature C (calc)_min | 35.53(2.38) | 35.53(2.4) | 0.21 | 0.83 |
| Arterial BP [Diastolic]_min | 33.46(13.95) | 33.49(14.18) | -0.15 | 0.88 |
| Non Invasive Blood Pressure systolic_min | 86.08(11.99) | 86.12(11.65) | -0.22 | 0.82 |
| Non Invasive Blood Pressure diastolic_min | 40.42(8.05) | 40.33(7.97) | 0.93 | 0.35 |
| Temperature Fahrenheit_min | 94.02(8.23) | 93.98(8.43) | 1.13 | 0.26 |
| Arterial BP [Systolic]_max | 166.22(22.91) | 165.72(23.44) | 1.58 | 0.11 |
| NBP [Systolic]_max | 150.96(22.11) | 150.57(22.33) | 1.31 | 0.19 |
| Respiratory Rate Set_max | 16.46(4.72) | 16.42(4.71) | 0.71 | 0.48 |
| Temperature F_max | 100.32(1.95) | 100.36(2.07) | 0.71 | 0.48 |
| Arterial BP [Diastolic]_max | 92.73(20.80) | 92.39(20.59) | 1.12 | 0.23 |
| Heart Rate_max | 102.97(6.02) | 101.98(14.46) | 1.14 | 0.26 |
| Heart Rate_diff | 58.12(27.83) | 58.02(27.67) | 0.28 | 0.78 |
| NBP [Systolic]_diff | 69.82(36.85) | 69.79(36.73) | 0.06 | 0.95 |
| Respiratory Rate_diff | 25.36(11.35) | 25.16(11.34) | 1.29 | 0.2 |
| Temperature F_diff | 5.52(9.69) | 5.42(9.17) | 0.81 | 0.42 |
| Bicarbonate_max_value | 30.32(4.64) | 30.36(4.92) | -0.53 | 0.6 |
| Bicarbonate_min_value | 20.17(4.59) | 20.19(4.61) | -0.38 | 0.7 |
| Serum_Creatinine_min_value | 0.80(0.67) | 0.80(0.71) | 0.37 | 0.71 |
| Serum_Creatinine_diff | 69.63(312.66) | 66.18(336.43) | 0.78 | 0.43 |
| Serum_Potassium_Concentration_max_value | 12.13(17.83) | 11.83(17.04) | 1.23 | 0.22 |
| Serum_Sodium_Concentration_min_value | 106.60(45.45) | 106.59(45.36) | 0.03 | 0.98 |
| Serum_Sodium_Concentration_diff | 38.07(47.77) | 38.21(49.47) | -0.19 | 0.85 |
| liver_failure_count | 0.07(0.33) | 0.07(0.32) | 0.5 | 0.62 |
| Chronic_heart_failure_count | 0.14(0.56) | 0.12(0.5) | 2.17 | 0.03 |
| organ_fail_count | 1.39(2.12) | 1.35(1.88) | 1.58 | 0.11 |
| sepsis_count | 0.15(0.44) | 0.15(0.43) | 0.56 | 0.58 |
| Uncomplicated_hypertension_count | 0.75(0.70) | 0.76(0.74) | -0.67 | 0.5 |
| Respiratory_dysfunction_count | 0.30(0.62) | 0.30(0.6) | 0.45 | 0.65 |

The results indicated that the training and testing sets were largely comparable. However, one variable, ‘Chronic heart failure count,’ reflected a significant difference between the datasets, as evidenced by its p-value. Overall, the analysis confirmed that, apart from this variable, there were no significant differences between the training and testing sets, ensuring that the model’s results are reliable and generalizable.

### Evaluation metrics proposed and baseline models’ performance

The results for both the proposed and baseline models are summarized in Table 3. The proposed approach, using the CatBoost model, achieved an AUROC of 0.862, an accuracy score of 0.789, and an F1 score of 0.747. These metrics highlight the model’s accuracy and robustness in reducing both Type I and Type II errors, validating its effectiveness for our predictive modeling tasks. The plot of the ROC curves for both the proposed and baseline models is presented in Fig 3.

**Table 3.** Summary of the evaluation metrics (AUCROC, Accuracy, Precision, Recall, F-Score) for the prediction models on the test set.

| Model | AUROC | Accuracy | Precision | Recall | F-Score |
| --- | --- | --- | --- | --- | --- |
| Decision Tree | 0.793 | 0.734 | 0.697 | 0.678 | 0.687 |
| Random Forest | 0.826 | 0.750 | 0.708 | 0.714 | 0.711 |
| KNN | 0.776 | 0.714 | 0.624 | 0.654 | 0.663 |
| Logistic Regression | 0.805 | 0.737 | 0.689 | 0.706 | 0.697 |
| SVM | 0.837 | 0.766 | 0.721 | 0.745 | 0.732 |
| XGBoost | 0.857 | 0.784 | 0.766 | 0.717 | 0.741 |
| CatBoost (Proposed) | 0.862 | 0.789 | 0.771 | 0.724 | 0.747 |

**Fig 3.**
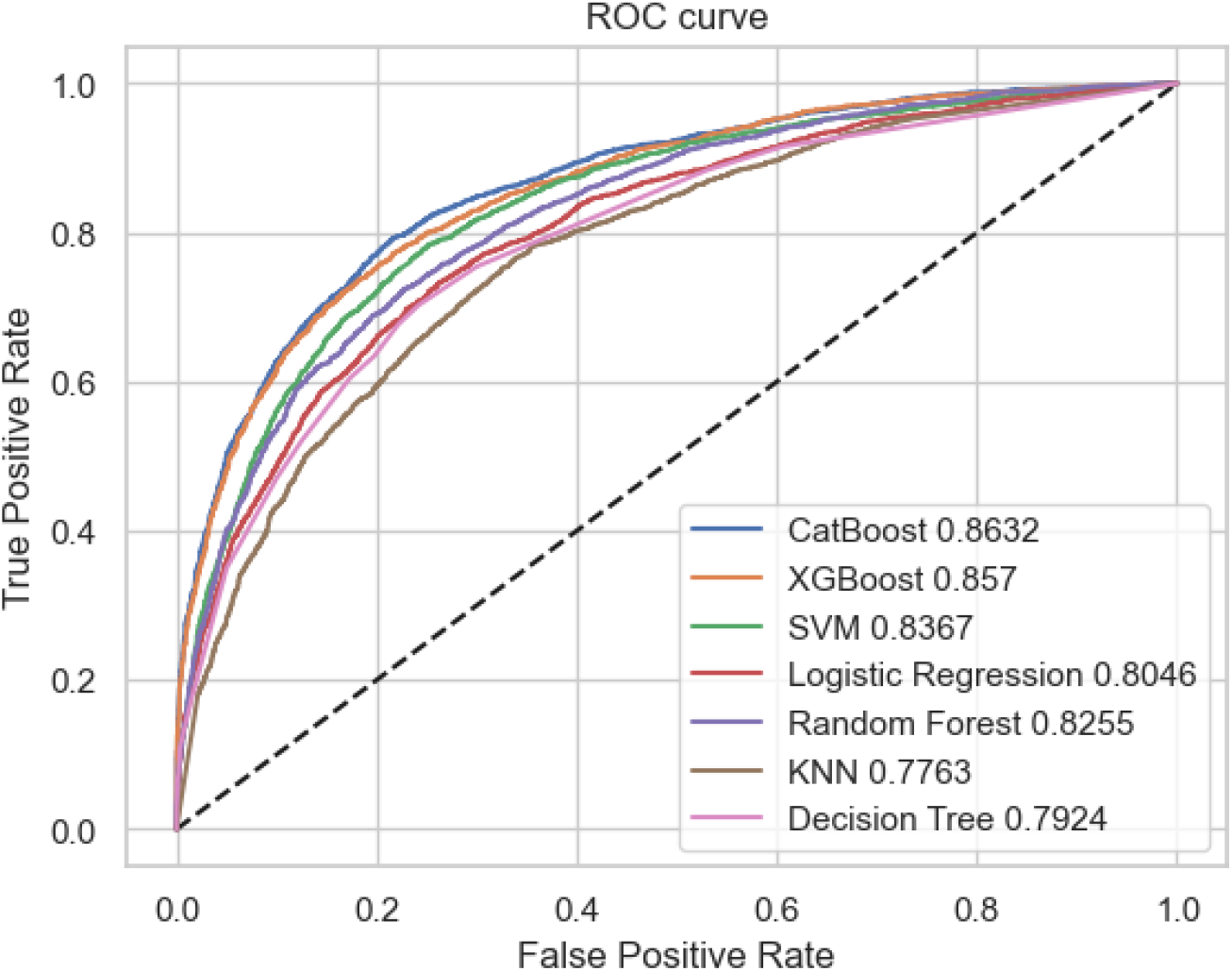
Model Comparison. The AUROC curves and scores for seven models: CatBoost, XGBoost, SVM, Logistic Regression, Random Forest, KNN, and Decision Tree.

Among the baseline models developed using the MIMIC-III database, the Support Vector Machine (SVM) and XGBoost models performed notably well, with high AUROC, accuracy, precision, recall, and F1 scores. In contrast, the Decision Tree and K-Nearest Neighbors (KNN) models showed relatively lower performance across these metrics. Overall, the CatBoost model emerged as the top-performing model, followed closely by SVM and XGBoost. The results indicate that the proposed approach outperforms the best baseline models.

Fig 4 shows the calibration curves for seven machine learning models: CatBoost, XGBoost, SVM, Logistic Regression, Random Forest, KNN, and Decision Tree. The X-axis represents the predicted probability, and the Y-axis represents the true probability, with the diagonal line indicating perfect calibration. Observations reveal that CatBoost, XGBoost, and Logistic Regression exhibit good calibration, closely aligning with the diagonal line. SVM and Random Forest show moderate calibration, with some deviation. In contrast, KNN and Decision Tree exhibit poor calibration, slightly deviating from the diagonal line. These results suggest that CatBoost, XGBoost, and Logistic Regression are better at predicting accurate probabilities.

**Fig 4.**
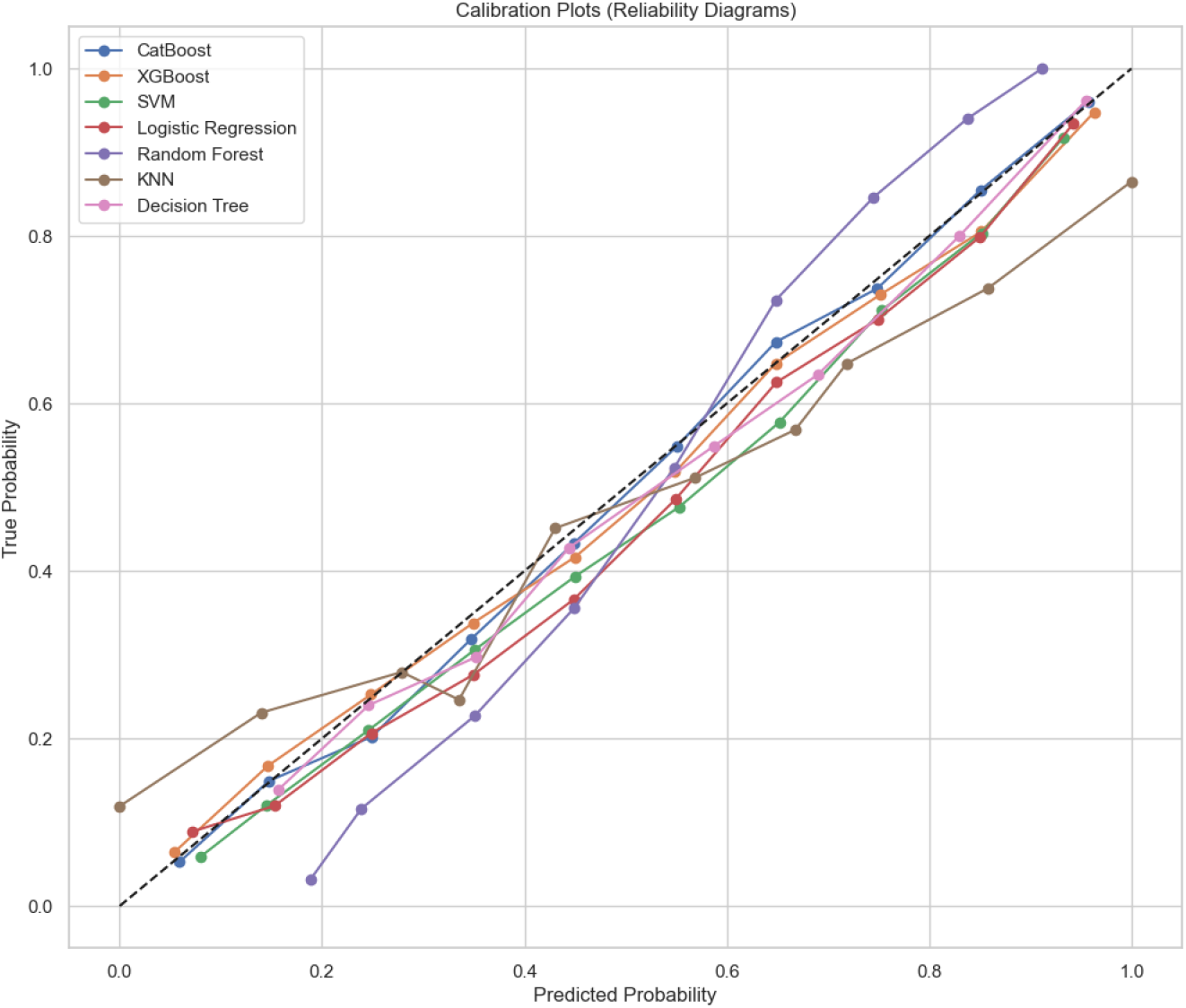
Calibration Curves for seven machine learning models, comparing predicted probabilities with true probabilities. CatBoost, XGBoost, and Logistic Regression show better calibration, while KNN and Decision Tree show poorer calibration.

## Discussion

### Summary of existing model compilation

Several models have been concurrently developed to predict mortality for ICU patients, specifically those receiving mechanical ventilation (MV). These include machine learning (ML) frameworks using various techniques and databases, with a particular focus on the MIMIC-III database due to its comprehensive nature. Notably, Zhu et al. utilized a wide range of features and ML methods, ultimately finding that the XGBoost model performed best with the highest AUC among their tested models.

In Zhu’s study, which also utilized the MIMIC-III database, data preprocessing resulted in a dataset of 25,659 patients and 55 predictors. Their features included demographics, ICU diagnosis, pre-ICU comorbidities, vital signs, disease severity scores, and laboratory test results from the first day of ICU admission. Various machine learning methods were employed, including KNN, bagging, logistic regression, decision tree, XGBoost, random forest, and neural networks [7].

In contrast to the existing literature, our study employs a more streamlined feature selection process, using only 32 features derived from backward elimination, the Lasso method, and expert clinical insights. We addressed class imbalances using the Synthetic Minority Over-sampling Technique (SMOTE) and managed missing data with median imputation and categorical transformations via Label Encoder. Our proposed approach leverages the CatBoost model, known for its superior handling of categorical variables and computational efficiency. The CatBoost model in our study achieved an AUROC of 0.862, a significant improvement from the initial 0.821 AUROC score, and demonstrated notable performance in terms of accuracy (0.789) and F1-score (0.747). This is a marked improvement over Zhu et al.’s findings, which, despite using more features (55 in total), did not achieve as high an AUROC score. Additionally, the calibration plots showed that our models are well-calibrated.

The key advantages of our approach over previous studies are multifold: (1) our systematic feature selection process, combined with clinical insights, ensures that only the most pertinent features are included, reducing complexity and enhancing model accuracy; (2) the application of SMOTE effectively addresses the imbalance in target variables, which was not considered in previous studies; and (3) CatBoost’s gradient boosting architecture and efficiency in handling high-dimensional data make it a superior choice for this application, outperforming traditional models in both accuracy and computational speed.

### Study limitations and future research

Our study has several limitations that need to be acknowledged. Firstly, we were unable to validate our model using external datasets due to the lack of access to comprehensive databases similar to MIMIC-III. This limitation restricts our ability to confirm the generalizability and robustness of our proposed model across different patient populations and clinical settings. Future research should focus on validating our models using external datasets to ensure their applicability in diverse healthcare environments.

Secondly, the MIMIC-III database we utilized is over ten years old and lacks comprehensive historical information for many patients. This could introduce bias in our dataset selection, potentially affecting the model’s performance and its relevance to current clinical practices. The reliance on an older database may not fully capture the advancements in ICU care and patient management that have occurred over the past decade. To address this limitation, future research should aim to leverage newer databases that reflect the latest clinical data and practices, thereby improving the model’s accuracy and applicability.

Additionally, while our study implemented advanced feature selection and data preprocessing techniques, there is still room for improvement in handling missing data and imbalances in the dataset. Future studies could explore more sophisticated imputation methods and balancing techniques to further enhance model performance. By addressing these limitations, future research can build on our findings and contribute to more robust and generalizable predictive models for ICU patient outcomes.

Future research should explore the integration of deep learning approaches, such as Convolutional Neural Networks (CNNs) and Recurrent Neural Networks (RNNs), which can provide significant improvements in handling complex and high-dimensional data. These methods are particularly useful for capturing temporal patterns and dependencies in the data, which are critical in predicting outcomes for ICU patients. Assessing the feasibility and effectiveness of these advanced models in the context of ICU mortality prediction could lead to more accurate and reliable predictions.

Furthermore, it is essential to consider the clinical implementation of predictive models. Developing user-friendly tools and interfaces that can be seamlessly integrated into clinical workflows will be crucial. Engaging with clinicians during the development process to understand their needs and preferences will ensure that the models are not only accurate but also practical and easy to use in everyday clinical practice. This approach will ultimately enhance the adoption and impact of predictive models in improving patient outcomes in the ICU.

Lastly, future studies should investigate the potential of employing real-time data streams and continuous learning models to keep the predictive systems up-to-date with the latest clinical data and practices. This dynamic approach could significantly improve the model’s responsiveness and accuracy, ensuring that it remains relevant and effective in rapidly evolving clinical environments.

## Conclusion

This study significantly improves the prediction of hospital mortality for ICU patients on mechanical ventilation using advanced machine learning techniques. By applying data imputation, bootstrapping, and model optimization, we enhanced our models’ predictive accuracy and robustness, as shown by the substantial improvements in AUC and accuracy metrics.

The CatBoost model, noted for its efficient handling of categorical variables and computational demands, proved particularly effective. Our streamlined feature selection process and the use of SMOTE ensured the inclusion of relevant features, reducing complexity and boosting model performance. Additionally, our models demonstrated good calibration, ensuring that predicted probabilities closely matched actual outcomes.

However, limitations remain. The absence of external dataset validation and reliance on the older MIMIC-III database may introduce biases and limit generalizability. Future research should validate our models with newer, diverse datasets to enhance applicability across clinical settings. Incorporating additional data types, such as clinical notes and images from the MIMIC-III dataset, could further improve model accuracy and reliability. Leveraging these data sources aims to develop models that perform well both in controlled environments and real-world clinical settings.

Ongoing research and integration of varied datasets and advanced data types will continue to improve model generalizability and performance, ultimately aiding clinicians in making informed decisions and improving patient outcomes in ICUs.

## Data Availability

All data produced are available online at https://physionet.org/content/mimiciii/1.4/

## Acknowledgments

The authors express their appreciation to the developers of MIMIC-III for providing a comprehensive and detailed public EHR dataset.

## Notes

### Competing Interest Statement

The authors have declared no competing interest.

### Funding Statement

This study did not receive any funding

## References

1. Kempker JA, Abril MK, Chen Y, Kramer MR, Waller LA, Martin GS. The epidemiology of respiratory failure in the United States 2002–2017: a serial cross-sectional study. Crit Care Explor. 2020;2:e0128.

2. Wunsch H, Wagner J, Herlim M, Chong DH, Kramer AA, Halpern SD. Occupancy and mechanical ventilator use in the United States. Crit Care Med. 2013;41:2712–2719.

3. Mehta AB, Syeda SN, Wiener RS, Walkey AJ. Epidemiological trends in invasive mechanical ventilation in the United States: A population-based study. J Crit Care. 2015;30(6):1250–1256.

4. Johnson A, Pollard T, Mark R. MIMIC-III Clinical Database (version 1.4). PhysioNet. 2016.

5. Gao J, Lu Y, Ashrafi N, Domingo I, Alaei K, Pishgar M. Prediction of Sepsis Mortality in ICU Patients Using Machine Learning Methods. medRxiv. 2024;2024.03.14.24304184.

6. Zhang J, Li H, Ashrafi N, Yu Z, Placencia G, Pishgar M. Prediction of In-Hospital Mortality for ICU Patients with Heart Failure. medRxiv. 2024;2024.06.25.24309448.

7. Zhu Y, Zhang J, Wang G, Yao R, Ren C, Chen G, et al. Machine Learning Prediction Models for Mechanically Ventilated Patients: Analyses of the MIMIC-III Database. Front Med. 2021;8:662340.

8. Su L, Zhang Z, Zheng F, et al. Five novel clinical phenotypes for critically ill patients with mechanical ventilation in intensive care units: a retrospective and multi database study. Respir Res. 2020;21:325.

9. Liang Y, Zhu C, Tian C, et al. Early prediction of ventilator-associated pneumonia in critical care patients: a machine learning model. BMC Pulm Med. 2022;22:250.

10. Yu L, Halalau A, Dalal B, Abbas AE, Ivascu FA, Amin M, et al. Machine learning methods to predict mechanical ventilation and mortality in patients with COVID-19. PLoS One. 2021;16(4):e0249285.

11. Yao RQ, Jin X, Wang GW, Yu Y, Wu GS, Zhu YB, et al. A machine learning-based prediction of hospital mortality in patients with postoperative sepsis. Front Med (Lausanne). 2020;7:445.

12. Hsu PC, Lin YT, Kao KC, et al. Risk factors for prolonged mechanical ventilation in critically ill patients with influenza-related acute respiratory distress syndrome. Respir Res. 2024;25:9.

13. Dai Z, et al. Analysis of adult disease characteristics and mortality on MIMIC-III. PLoS One. 2020;15(4):e0232176.

14. Lin Z, Huang X, Shan X. Development and validation of a survival prediction model for patients received mechanical ventilation in the intensive care unit: a large sample size cohort from the MIMIC database. Ann Palliat Med. 2022;11(6):2071–2084.

15. Sayed M, Rian∼o D, Villar J. Predicting Duration of Mechanical Ventilation in Acute Respiratory Distress Syndrome Using Supervised Machine Learning. J Clin Med. 2021;10(17):3824.

16. Li L, Zhang Z, Xiong Y, Hu Z, Liu S, Tu B, Yao Y. Prediction of hospital mortality in mechanically ventilated patients with congestive heart failure using machine learning approaches. Int J Cardiol. 2022; Version of Record 16 May 2022.

17. Van Calster B, Wynants L, Timmerman D, Steyerberg EW, Collins GS. Predictive analytics in healthcare: how can we know it works? J Am Med Inform Assoc. 2019;26(12):1651–1654.

18. Harutyunyan H, Khachatrian H, Kale DC, Ver Steeg G, Galstyan A. Multitask learning and benchmarking with clinical time series data. Sci Data. 2019;6:96.

19. George N, Moseley E, Eber R, Siu J, Samuel M, Yam J, et al. Deep learning to predict long-term mortality in patients requiring 7 days of mechanical ventilation. PLoS One. 2021;16(6):e0253443.

20. Sauer CM, Sasson D, Paik KE, McCague N, Celi LA, Sánchez Fernández I, et al. Feature selection and prediction of treatment failure in tuberculosis. PLoS One. 2018;13(11):e0207491.

21. van Egmond MB, Spini G, van der Galien O, IJpma A, Veugen T, Kraaij W, et al. Privacy-preserving dataset combination and Lasso regression for healthcare predictions. BMC Med Inform Decis Mak. 2021;21(1):266.

22. Elreedy D, Atiya AF. A comprehensive analysis of synthetic minority oversampling technique (SMOTE) for handling class imbalance. Inf Sci. 2019;505:32–64.

23. Parodi S, Verda D, Bagnasco F, Muselli M. The clinical meaning of the area under a receiver operating characteristic curve for the evaluation of the performance of disease markers. Epidemiol Health. 2022;44.

24. Prokhorenkova L, Gusev G, Vorobev A, Dorogush AV, Gulin A. CatBoost: unbiased boosting with categorical features. Adv Neural Inf Process Syst. 2018;31:6638–6648.

25. Seto H, Oyama A, Kitora S, et al. Gradient boosting decision tree becomes more reliable than logistic regression in predicting probability for diabetes with big data. Sci Rep. 2022;12:15889.

26. Mbonyinshuti F, Nkurunziza J, Niyobuhungiro J, Kayitare E. Application of random forest model to predict the demand of essential medicines for non-communicable diseases management in public health facilities. Pan Afr Med J. 2022;42:89.

27. Zhou X, Li X, Zhang Z, et al. Support vector machine deep mining of electronic medical records to predict the prognosis of severe acute myocardial infarction. Front Physiol. 2022;13:991990.

28. Xing W, Bei Y. Medical Health Big Data Classification Based on KNN Classification Algorithm. IEEE Access. 2019.

29. Schober P, Vetter TR. Logistic Regression in Medical Research. Anesth Analg. 2021;132(2):365–366.

30. Zhang Z, Zhao Y, Canes A, Steinberg D, Lyashevska O. Predictive analytics with gradient boosting in clinical medicine. Ann Transl Med. 2019;7(7):152.

31. Aliya A, Yurf Asghar S, Danish Khan Yousafzai A, Haider Bangash A, Mohsin R, Fatima A, et al. Prediction of In-Hospital Mortality Among Heart Failure Patients: An Automated Machine Learning Analysis of Mimic-III Database. Am Heart J. 2022;254:261.

32. Safaei N, Safaei B, Seyedekrami S, Talafidaryani M, Masoud A, Wang S, et al. E-CatBoost: An efficient machine learning framework for predicting ICU mortality using the eICU Collaborative Research Database. PLoS One. 2022;17(5):e0262895.

33. Demsar J. Statistical Comparisons of Classifiers over Multiple Data Sets. J Mach Learn Res. 2006;7:1–30.

34. Nohara Y, Matsumoto K, Soejima H, Nakashima N. Explanation of machine learning models using Shapley additive explanation and application for real data in hospital. Comput Methods Programs Biomed. 2022;214:106584.

